# Prediction and Characterization of Genetically Regulated Expression of Target Tissues in Asthma

**DOI:** 10.1101/2025.02.06.25321273

**Authors:** Sarah D. Slack, Erika Esquinca, Christopher H. Arehart, Meher Preethi Boorgula, Brooke Szczesny, Alex Romero, Monica Campbell, Sameer Chavan, Nicholas Rafaels, Harold Watson, R. Clive Landis, Nadia N. Hansel, Charles N. Rotimi, Christopher O. Olopade, Camila A. Figueiredo, Carole Ober, Andrew H. Liu, Eimear E. Kenny, Kai Kammers, Ingo Ruczinski, Margaret A. Taub, Michelle Daya, Christopher R. Gignoux, Katerina Kechris, Kathleen C. Barnes, Rasika A. Mathias, Randi K. Johnson

## Abstract

**Background:** Genetic control of gene expression in asthma-related tissues is not well-characterized, particularly for African-ancestry populations, limiting advancement in our understanding of the increased prevalence and severity of asthma in those populations.

**Objective:** To create novel transcriptome prediction models for asthma tissues (nasal epithelium and CD4+ T cells) and apply them in transcriptome-wide association study (TWAS) to discover candidate asthma genes.

**Methods:** We developed and validated gene expression prediction databases for unstimulated CD4+ T cells (CD4+T) and nasal epithelium using an elastic net framework. Combining these with existing prediction databases (N=51), we performed TWAS of 9,284 individuals of African-ancestry to identify tissue-specific and cross-tissue candidate genes for asthma. For detailed Methods, please see the Supplemental Methods.

**Results:** Novel databases for CD4+T and nasal epithelial gene expression prediction contain 8,351 and 10,296 genes, respectively, including four asthma loci (*SCGB1A1, MUC5AC, ZNF366, LTC4S*) not predictable with existing public databases. Prediction performance was comparable to existing databases and was most accurate for populations sharing ancestry with the training set (e.g. African ancestry). From TWAS, we identified 17 candidate causal asthma genes (adjusted *P*<0.1), including genes with tissue-specific (*IL33* in nasal epithelium) and cross-tissue (*CCNC* and *FBXW7*) effects.

**Conclusions:** Expression of *IL33, CCNC*, and *FBXW7* may affect asthma risk in African ancestry populations by mediating inflammatory responses. The addition of CD4+T and nasal epithelium prediction databases to the public sphere will improve ancestry representation and power to detect novel gene-trait associations from TWAS.

**Key Messages:**

- From the largest African-ancestry TWAS of asthma to date (N=9,284), we identified 17 candidate causal asthma genes, including: nasal epithelial expression of *IL33*, and cross-tissue expression of *CCNC* and *FBXW7*.
- We provide gene expression prediction databases for CD4+ T cell and nasal epithelial tissues built in African-ancestry populations, improving ancestry representation and power to detect novel gene-trait associations from TWAS.

**Capsule Summary:** We developed novel gene expression prediction databases (CD4+ T cells, nasal airway epithelium) representing diverse populations across the African diaspora and identified 17 candidate causal asthma genes from TWAS.

## INTRODUCTION

Identification of the genetic control of gene expression through expression quantitative trait locus (eQTL) mapping has improved our understanding of the genetic basis for human disease, including asthma(1). Efforts to catalogue eQTLs have enabled large-scale transcriptome-wide association studies (TWAS) leveraging widely available genetic variation to predict gene expression and test for association with phenotypes(2). The accuracy of these imputations are directly influenced by the underlying eQTL architecture(3) which varies across tissue and by ancestry(4). Given the vast over-representation in genomic studies of individuals of European descent, existing reference transcriptome datasets were trained primarily using European ancestry populations, resulting in poor cross-ancestry prediction quality(3–6). To date, only two reference transcription prediction databases are publicly available for populations of African ancestry: monocytes(4) and whole blood(7).

CD4+ T cells (CD4+T) and the nasal airway epithelium (nasal epithelium) both play a central role in modulating allergic disease and asthma. Allergens induce a CD4+T helper cell response, which drives airway inflammation through type 2 cytokines. The airway epithelium plays critical roles in inflammation, leading to barrier dysfunction and airway remodeling in asthma. However, expression in these tissues is not well-characterized in public repositories (e.g. GTEx), particularly for African-ancestry populations that are disproportionately affected by the disease(8) and have distinct genetic risk factors(9, 10). Identification of expression signatures associated with asthma in these target tissues can give new insight into the genetics driving dysfunction in allergic disease.

We aimed to overcome these gaps in tissue and ancestry representation for eQTL and TWAS using data from African-ancestry populations in the Consortium on Asthma among African-ancestry Populations in the Americas (CAAPA). We built predictive models to estimate genetically-driven gene expression in the nasal epithelium and for CD4+T, evaluated model accuracy and gene contents, and then applied both the novel and existing prediction databases in TWAS to identify candidate asthma genes (**Figure 1**).

**Figure 1.**
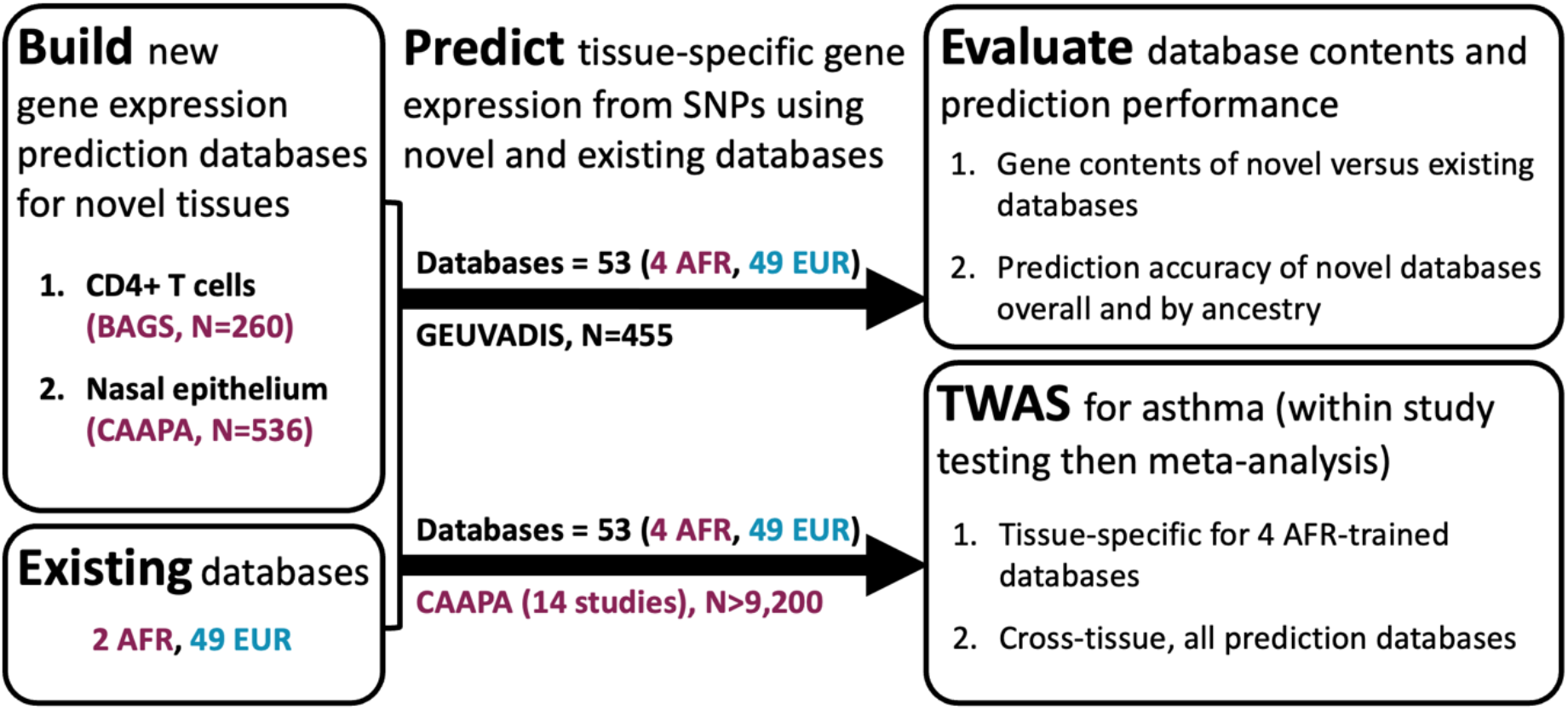
Study overview. Colors indicate predominant global ancestry as African (AFR, purple) or European (EUR, blue) for each study population or prediction database.

## RESULTS AND DISCUSSION

### Gene Expression Prediction Databases and Accuracy

We developed gene expression prediction databases for unstimulated CD4+T (N=260) and nasal epithelium (N=536) using RNA sequencing (RNAseq) and genome-wide association study (GWAS) data (**Table E1**). CD4+T RNAseq was available from Barbados Asthma Genetics Study (BAGS) participants only, while nasal epithelium RNAseq represented BAGS and six other CAAPA studies(11). The Yoruban (YRI) ancestry ranged from 0.09 to 0.99 (median=0.88) for BAGS and 0.09 to 0.99 (median=0.84) for CAAPA (**Figure E1**). Genes achieving significant prediction models (model-based prediction accuracy R^2^>0.01 and *P*<0.05) were included in prediction databases. The R^2^ from nested cross-validation ranged from 0.01 to 0.75 (N=8,351, median=0.12) for CD4+T and 0.01 to 0.80 (N=10,296, median=0.12) for nasal epithelium, which is comparable with existing databases for monocytes(4), whole blood(2), and lung(2) tissues.

We evaluated the accuracy of the novel prediction databases overall and across continental ancestral populations using the GEUVADIS dataset(12). Prediction accuracy calculated in all GEUVADIS populations combined ranged from 0 to 0.83 in CD4+T and 0 to 0.81 in nasal epithelium. When calculated separately, accuracy was significantly different by population (Kruskal-Wallis *P*<0.05) for genes with model prediction accuracy in the top 40% (**Figure 2A**), in particular for CD4+T. The asthma candidate gene *GSTM1* exemplifies this pattern of predictive accuracy across subpopulations (**Figure 2B**). Consistent with prior reports(3–6), predictive accuracy from these CD4+T and nasal epithelium models is higher in populations with increased ancestral similarity to the training datasets.

**Figure 2.**
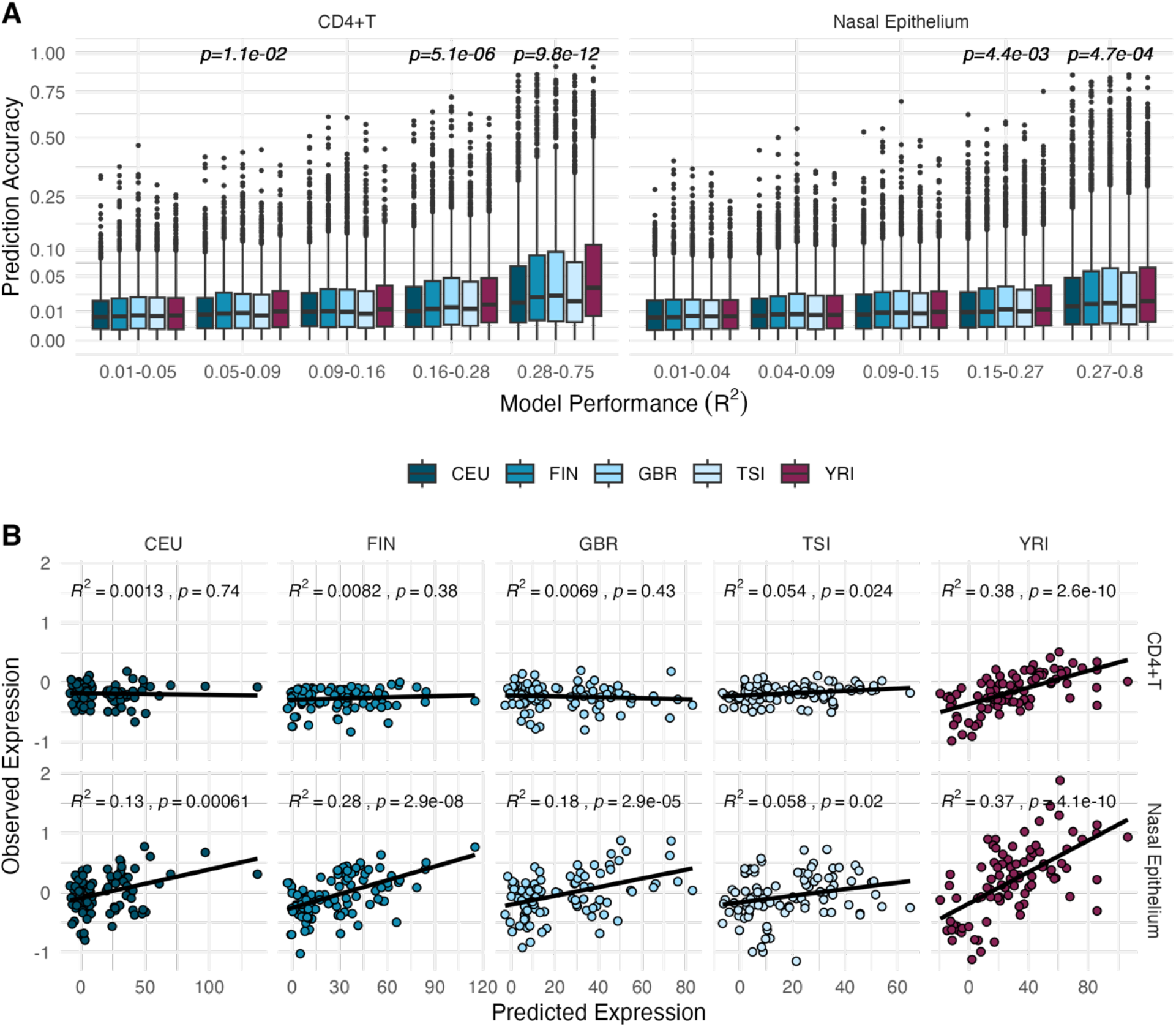
Prediction accuracy of novel CD4+T and nasal epithelium databases in GEUVADIS. A) Accuracy of predicted versus observed expression within model performance quintiles, with Kruskal-Wallis test of population differences. B) Pearson correlation of observed versus predicted expression for candidate asthma gene *GSTM1. CEU = Utah residents, FIN = Finnish, GBR = British, TSI = Toscani, Italy, YRI = Yoruba*.

We examined the overlap between genes in the novel databases with a) existing publicly available prediction databases, and b) 172 known asthma and allergy candidates from a recent review(13) (**Figure E2**). More (>50%) asthma and allergy candidate genes were well-predicted in the nasal epithelium compared to CD4+T (30%). Multiple candidate genes not predictable in currently available databases (N=51, see “Prediction of Gene Expression” in the Supplemental Methods) were newly predicted, including *SCGB1A1* (R^2^=0.09) and *MUC5AC* (R^2^=0.17). In total, 217 genes were newly predicted in CD4+T and nasal epithelium compared to existing databases(4, 7, 14). The 121 genes predicted for the first time in the nasal epithelium were enriched for genes involved in the structural constituent of chromatin (GO:0030527, *P*=1.08×10-5), see the Supplemental Methods). Genes newly predicted in CD4+T were not enriched for any gene sets.

### Asthma Transcriptome-Wide Association Study (TWAS)

We performed TWAS using PrediXcan(2) and MultiXcan(15) frameworks to identify tissue-specific and cross-tissue (respectively) candidate genes for asthma. These frameworks leverage SNP genotypes weighted by prediction model databases to predict gene expression and perform gene-based association testing to identify candidate causal genes. The study populations included 9,284 CAAPA participants (excluding individuals used in prediction model building, see the Supplemental Methods), representing genetic diversity across the African Diaspora at 14 studies/sites (YRI ancestry=34-88%, **Table E2**).

We performed tissue-specific TWAS in each study using predicted expression from the databases trained in African-ancestry populations: CD4+T, nasal epithelium, monocytes(4) and whole blood(7). We meta-analyzed results across studies by tissue, resulting in four tissue-specific TWAS. After multiple comparison correction of meta-analysis p-values, we identified three genes associated with asthma in CAAPA (adjusted *P*<0.1, **Figure 3A, Table E3**), each from a different tissue TWAS: *IL33* (nasal epithelium), *ST8SIA6* (whole blood), and *CSRP1* (monocytes). Participants with asthma had increased expression of *IL33* in nasal epithelium, *ST8SIA6* in whole blood, and *CSRP1* in monocytes compared to controls and independent of age, sex, kinship, and genetic ancestry (**Figure 3A, Figure E3**).

**Figure 3.**
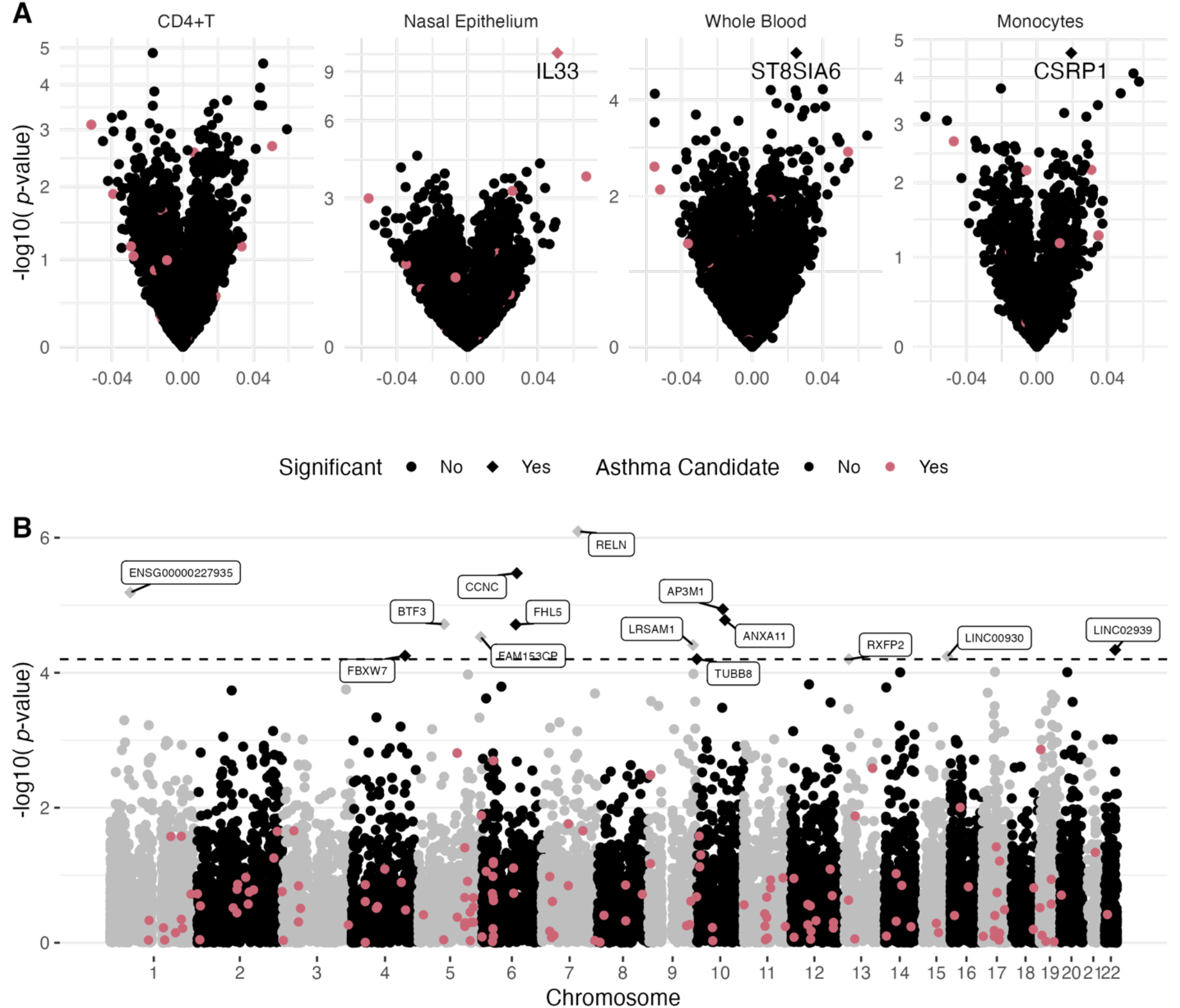
A) Meta-analysis of tissue-specific TWAS for asthma, where predicted expression of labeled genes marked by diamonds met the FDR-adjusted *P*<0.1 threshold for significance, set separately within each tissue. B) Meta-analysis of cross-tissue MultiXcan TWAS for asthma. Predicted expression of labeled genes with diamond points was significantly associated with asthma (FDR-adjusted *P*<0.1).

The increased predicted expression of *IL33* in the nasal epithelium of cases is consistent with prior asthma GWAS(1, 10), TWAS(1, 16), and differential expression analyses(17, 18). IL33 is a master cytokine regulator that induces the type 2 inflammatory cascade characteristic in a majority of asthma cases(19). Nasal epithelium *IL33* prediction was most strongly influenced by variation in rs1888909, a putative causal regulatory variant where additional copies of the T allele confer increased risk for asthma through mediating increased mRNA expression and protein levels in nasal airway epithelial cells(20). This putative causal risk allele is more common in African ancestry populations (gnomAD allele frequency (AF)=0.48) compared to European (AF=0.25) or Admixed American (AF=0.23). We could not examine cross-tissue expression of *IL33* due to its limited presence in other tissue databases.

To evaluate shared tissue signal, we performed MultiXcan with all available databases (four African- and 51 European-ancestry trained)(4, 7, 14). We identified 14 genes significantly associated with asthma (FDR-adjusted *P*<0.1, **Figure 3B, Table E4**). The top genes identified by MultiXcan, *RELN* and *CCNC*, were recently identified through differential expression analysis of CAAPA nasal epithelium(11). The other 12 genes have not been associated with asthma previously: *ENSG00000227935, ANXA11, FHL5, AP3M1, BTF3, FAM153CP, LRSAM1, LINC02939, LINC00930, TUBB8, RXFP2, FBXW7*.

The multi-tissue associations for *CCNC* and *FBXW7* represent broad generalizability across tissues and diverse ancestries. Nine tissues contributed to the *CCNC* PC-regression, and 10 tissues contributed to *FBXW7* (**Table E4**). The physical interaction of the *CCNC*-encoded cyclin D protein with CDK8(21), a transcription-regulating kinase, is essential for activating its inflammatory mediating response(21). These cyclin-dependent kinases may be promising therapeutic targets for asthma(22). *FBXW7* may also influence asthma through mediating airway remodeling or eosinophilic inflammation. Variants in *FBXW7* associate with eosinophil counts in trans-ancestry GWAS of blood cell traits(23). The F-box protein encoded by *FBXW7* is a key binding component of ubiquitin ligase complexes, acting through NOTCH1 and KLF5 targets to determine airway epithelial cell fates(24). Given their known biological functions and discovery in this genetic context, these candidates may contribute to reported ancestry differences in eosinophilic airway inflammation among asthma patients(25).

Major strengths of our TWAS include large sample size, diverse representation across the African diaspora, and inclusion of tissue-specific and cross-tissue discovery approaches. As a result, in the largest African-ancestry TWAS to date, we identified 17 candidate asthma genes for further investigation including *IL33, CCNC*, and *FBXW7*, of which 14 were novel and two have been identified only through CAAPA analyses*(11). IL33* was the only established asthma candidate identified and was consistent with prior nasal epithelium TWAS(16). Our inability to identify associations for other established asthma loci could reflect decreased power due to relatively lower sample size compared to other recent asthma TWAS (UK Biobank N>300,000(1, 16)), inability to distinguish between adult-onset and child-onset asthma in our TWAS populations, or may reflect consistency with prior evidence for distinct genetic risk factors for asthma in African-ancestry populations(10).

We developed publicly available [Zenodo link] gene expression prediction databases for two novel tissues implicated in asthma (CD4+T, nasal airway epithelium) and trained using diverse populations across the African diaspora. These databases allow for expression prediction of some candidate asthma genes (e.g., *MUC5AC*) for the first time. We add these resources to existing prediction databases trained in European-(N=49) and African-(N=2) ancestry populations. Shared ancestry in prediction training and testing datasets is an established factor affecting prediction accuracy and TWAS statistical power, second only to sample size(3, 4).

Incorporating ancestrally diverse training models has been shown to increase TWAS gene-trait discoveries by as much as 78%(7). By doubling the available expression prediction databases trained with admixed populations, not only do we improve representation of populations with the highest prevalence of and the most severe asthma(8), we improve power for TWAS discovery across a multitude of traits.

## Data Availability

All data produced will be available online at the time of publication.

## Abbreviations

BAGS: Barbados Asthma Genetics Study
CAAPA: Consortium on Asthma among African-ancestry Populations in the Americas
eQTL: expression quantitative trait locus
TWAS: transcriptome-wide association study
GWAS: genome-wide association study
RNAseq: RNA sequencing

## Acknowledgements

The authors would like to thank the participants at all CAAPA sites for their involvement and sample donation, as well as the staff who facilitated sample collection. The authors also wish to acknowledge the contributions of the consortium working on the development of the NHLBI BioData Catalyst® (BDC) ecosystem. This work was supported in part by the Division of Intramural Research, National Institute of Allergy and Infectious Diseases, NIH.

## REFERENCES

1. Pividori M, Schoettler N, Nicolae DL, Ober C, Im HK. Shared and distinct genetic risk factors for childhood-onset and adult-onset asthma: genome-wide and transcriptome-wide studies. Lancet Respir Med. 2019;7(6):509–22.

2. Gamazon ER, Wheeler HE, Shah KP, Mozaffari SV, Aquino-Michaels K, Carroll RJ, et al. A gene-based association method for mapping traits using reference transcriptome data. Nat Genet. 2015;47(9):1091–8.

3. Keys KL, Mak ACY, White MJ, Eckalbar WL, Dahl AW, Mefford J, et al. On the cross-population generalizability of gene expression prediction models. PLOS Genetics. 2020;16(8):e1008927.

4. Mogil LS, Andaleon A, Badalamenti A, Dickinson SP, Guo X, Rotter JI, et al. Genetic architecture of gene expression traits across diverse populations. PLoS Genet. 2018;14(8):e1007586.

5. Geoffroy E, Gregga I, Wheeler HE. Population-Matched Transcriptome Prediction Increases TWAS Discovery and Replication Rate. iScience. 2020;23(12):101850.

6. Mikhaylova AV, Thornton TA. Accuracy of Gene Expression Prediction From Genotype Data With PrediXcan Varies Across and Within Continental Populations. Front Genet. 2019;10:261.

7. Kachuri L, Mak ACY, Hu D, Eng C, Huntsman S, Elhawary JR, et al. Gene expression in African Americans, Puerto Ricans and Mexican Americans reveals ancestry-specific patterns of genetic architecture. Nature Genetics. 2023.

8. Moorman JE, Akinbami LJ, Bailey C, Zahran H, King M, Johnson C, et al. National surveillance of asthma: United States, 2001-2010. Vital & health statistics Series 3, Analytical and epidemiological studies. 2012(35):1–58.

9. Daya M, Rafaels N, Brunetti TM, Chavan S, Levin AM, Shetty A, et al. Association study in African-admixed populations across the Americas recapitulates asthma risk loci in non-African populations. Nat Commun. 2019;10(1):880.

10. Chang X, March M, Mentch F, Qu H, Liu Y, Glessner J, et al. Genetic architecture of asthma in African American patients. J Allergy Clin Immunol. 2023;151(4):1132–6.

11. Szczesny B, Boorgula MP, Chavan S, Campbell M, Johnson RK, Kammers K, et al. Multi-omics in nasal epithelium reveals three axes of dysregulation for asthma risk in the African Diaspora populations. Nat Commun. 2024;15(1):4546.

12. Lappalainen T, Sammeth M, Friedländer MR, t Hoen PA, Monlong J, Rivas MA, et al. Transcriptome and genome sequencing uncovers functional variation in humans. Nature. 2013;501(7468):506–11.

13. Kanchan K, Clay S, Irizar H, Bunyavanich S, Mathias RA. Current insights into the genetics of food allergy. J Allergy Clin Immunol. 2021;147(1):15–28.

14. Barbeira AN, Bonazzola R, Gamazon ER, Liang Y, Park Y, Kim-Hellmuth S, et al. Exploiting the GTEx resources to decipher the mechanisms at GWAS loci. Genome Biol. 2021;22(1):49.

15. Barbeira AN, Pividori M, Zheng J, Wheeler HE, Nicolae DL, Im HK. Integrating predicted transcriptome from multiple tissues improves association detection. PLoS Genet. 2019;15(1):e1007889.

16. Sajuthi SP, Everman JL, Jackson ND, Saef B, Rios CL, Moore CM, et al. Nasal airway transcriptome-wide association study of asthma reveals genetically driven mucus pathobiology. Nat Commun. 2022;13(1):1632.

17. Forno E, Zhang R, Jiang Y, Kim S, Yan Q, Ren Z, et al. Transcriptome-wide and differential expression network analyses of childhood asthma in nasal epithelium. J Allergy Clin Immunol. 2020;146(3):671–5.

18. Tsai YH, Parker JS, Yang IV, Kelada SNP. Meta-analysis of airway epithelium gene expression in asthma. Eur Respir J. 2018;51(5).

19. Fahy JV. Type 2 inflammation in asthma--present in most, absent in many. Nat Rev Immunol. 2015;15(1):57–65.

20. Aneas I, Decker DC, Howard CL, Sobreira DR, Sakabe NJ, Blaine KM, et al. Asthma-associated genetic variants induce IL33 differential expression through an enhancer-blocking regulatory region. Nat Commun. 2021;12(1):6115.

21. Knuesel MT, Meyer KD, Donner AJ, Espinosa JM, Taatjes DJ. The human CDK8 subcomplex is a histone kinase that requires Med12 for activity and can function independently of mediator. Mol Cell Biol. 2009;29(3):650–61.

22. Barnes PJ. Kinases as Novel Therapeutic Targets in Asthma and Chronic Obstructive Pulmonary Disease. Pharmacol Rev. 2016;68(3):788–815.

23. Chen MH, Raffield LM, Mousas A, Sakaue S, Huffman JE, Moscati A, et al. Trans-ethnic and Ancestry-Specific Blood-Cell Genetics in 746,667 Individuals from 5 Global Populations. Cell. 2020;182(5):1198–213.e14.

24. Li R, Zhang Y, Garg A, Sui P, Sun X. E3 ubiquitin ligase FBXW7 balances airway cell fates. Dev Biol. 2022;483:89–97.

25. Nyenhuis SM, Krishnan JA, Berry A, Calhoun WJ, Chinchilli VM, Engle L, et al. Race is associated with differences in airway inflammation in patients with asthma. J Allergy Clin Immunol. 2017;140(1):257–65.e11.

