## Supplemental Methods for "Prediction and Characterization of Genetically Regulated Expression of Target Tissues in Asthma"

#### Study Populations

*The Consortium on Asthma among African-Ancestry Populations in the Americas (CAAPA)* is a multi-national asthma study that has pioneered scientific efforts to characterize the distinct genetic architecture across the African Diaspora since 2011. More than 9,500 CAAPA participants have genotyping array data and asthma phenotyping, representing diverse geographic locations across the United States (Maryland, Illinois, Washington DC, North Carolina), Central (Honduras, Jamaica, Columbia, Barbados) and South America (Brazil). Participants in these studies were unrelated cases and controls, except for subjects from the Barbados Asthma Genetics Study (BAGS), which included both family-based and case-control recruitment, the Brazilian Immunogenetics of Asthma and Schistosomiasis (BIAS) study, where a whole-population ascertainment was used (including families), and the Howard University

Family Study (HUFS) where participants were a mixture of unrelated and related subjects. Definition of asthma was based on a doctor's diagnosis and/or standardized questionnaires; controls were defined as a reported negative history of asthma. Study-specific details for recruitment, phenotyping, and genotyping have been previously published[1]. Study characteristics of these 14 genetics studies, including geographic location, ancestry, genotyping technology, and asthma sample size, are provided in **Table E2**.

The CAAPA consortium also sought to understand multi-omics signatures for asthma and recruited a subset of participants (N=536) for nasal epithelial sampling from which RNA sequencing (RNAseq) data were generated and paired with MEGA genotyping array data for expression quantitative trait locus (eQTL) analyses. Participants represented seven geographic sites, including four US-based locations (Baltimore, Chicago, Denver, Washington D.C.) and three international locations (Barbados, Brazil, Nigeria). Detailed recruitment, data processing, harmonization, and discovery of differentially expressed genes distinguishing asthma cases from controls who never had asthma have been previously published[2]. All multi-omics samples were obtained following written informed consent from participants. The University of Colorado (IRB#: 17-1807), Johns Hopkins University (IRB00179053), University of Chicago (IRB18-0466-CR001), National Institutes of Health (IRB#: P184385), University of West Indies (IRB#: 190604-A), University of Bahia (IRB#: 3.302.487) and University of Ibadan (IRB18-0840) Institutional Review Boards approved the conduct of this study.

*The Barbados Asthma Genetics Study (BAGS)*, one of the studies involved in CAAPA, enrolled >1,600 asthma cases, family members, and unrelated controls through referrals at local polyclinics, the Accident and Emergency Department at the Queen Elizabeth Hospital and the Chronic Disease Research Centre since its inception in 1993. Asthma was defined based on a history of both self-reported and physician-diagnosed asthma, as well as a history of wheezing without an upper respiratory tract infection. Controls were selected based on no history of asthma. Nurse coordinators contacted and consented participants, and administered follow-up interviews to evaluate atopic asthma status[3] and asthma severity[4], including baseline lung function and collection of biospecimens. For the CD4+T RNAseq study, all subjects gave written consent as approved at Johns Hopkins, the University of West Indies, and the University of Colorado (COMIRB#: 15-2148).

*Genetic Deduplication.* Varying subsets of BAGS participants were included in the Trans-Omics for Precision Medicine (TOPMed) whole-genome sequencing (WGS) project (BAGS-WGS), the CD4+T eQTL project, and the CAAPA multi-omics project. Similarly, CAAPA participants could have been selected for both genome-wide association study (GWAS)[1] and multi-omics[2] study. Due to this overlapping recruitment, we used identity by descent analysis (IBD) in PLINK1.9[5] to identify genetic duplicates between analyses. After aligning all datasets to the TOPMed reference panel, we merged the common variants, completed (linkage disequilibrium (LD) pruning in PLINK with --indep-pairwise (10,000 variant count window, 5 variant step size, 0.3 $R^2$ threshold), finally calculating IBD in PLINK with --genome. Subject pairs were classified as duplicates if the Z2 (P(IBD=2)) value was greater than 0.97. To maximize transcriptome prediction sample size, duplicates with available RNAseq data (CD4+T or nasal epithelium) were retained in eQTL analyses and the duplicate removed from transcriptome-wide association studies (TWAS). For multi-tissue analysis with MultiXcan, deduplication was applied to affected CAAPA studies BAGS-WGS and ProAR before prediction with all tissue databases, and sample sizes across analysis populations represent non-overlapping participants after genetic deduplication (**Tables E1 and E2**). For single-tissue analysis, deduplication was only applied where necessary for BAGS-WGS and ProAR to preserve sample size where possible. BAGS-WGS sample size for prediction of all tissues (N=920) was consistent except for prediction of CD4+T (N=819) and nasal epithelium (N=895). ProAR sample size was consistent (N = 1107) for prediction of all tissues except CD4+T (N=1036).

**Genotyping and Whole-Genome Sequencing**

*Whole Genome Sequencing.* BAGS WGS was performed by Illumina in Phase I of the TOPMed initiative. We accessed BAGS WGS Freeze 8 data at dbGaP Study Accession phs001143.v4.p1 and filtered to only SNPs before Predixcan/Multixcan.

*Array Data.* CAAPA and BAGS (CD4+T) genotypes were generated from genomic DNA using various genome-wide array technologies, outlined in **Table E2**. For each study, we performed standard quality control procedures. We excluded samples that failed sex verification, heterozygosity checks, or had missingness > 5%. Participant deduplication was performed within and across studies as described above (see "Genetic Deduplication"). Genome-wide array data is available at dbGaP Study Accession: phs001123.v2.p1. We prepared the CAAPA and BAGS array data for PredictDB/Predixcan/Multixcan following the same quality control procedures. We filtered monoallelic SNPs, strand ambiguous SNPs, SNPs out of Hardy Weinberg equilibrium, and SNPs with missingness > 5% before imputing each study to the TOPMed r2 reference panel. SNPs with imputation $R^2>0.7$ and MAF>0.01 were retained for use in PredictDB/PrediXcan/MultiXcan.

**Nasal Epithelium RNA Sequencing**

Specimen collection and sequencing protocols have been published in detail[2]. Nasal ciliated epithelial cells from the posterior surface of the inferior turbinate were collected using cytology brushes and standardized protocols. DNA and RNA were extracted from the same nasal sample for multi-omics analysis. CAAPA RNAseq data is available in the GEO database under the accession code GSE240567 [https://www.ncbi.nlm.nih.gov/geo/query/acc.cgi?acc=GSM7701971]. We performed quality control (QC) pre-alignment, QC, adapter trimming, and alignment of reads to GRCh38 using FastQC[6], Picard[7], BBDuk[8], and HISAT2[9], respectively. Raw counts were generated by CoCo[10]. Full details are available in Szczesny et al[2].

**CD4+T Cell RNA Sequencing**

*Sample Collection and Sequencing.* Venous blood was collected by simple venipuncture under aseptic conditions. All samples were processed within two hours of collection to minimize gene expression variations associated with longer sample incubation times. Peripheral blood mononuclear cells (PBMCs) were separated by the Ficoll density gradient method and immediately processed to isolate CD4+ T cells (CD4+T). Using the CD4+ T Cell Isolation Kit (Miltenyi Biotec Inc), human CD4+ T helper cells were isolated from PBMCs, by depletion of non-target cells (negative selection). Non-target cells were labeled with a cocktail of biotin-conjugated monoclonal antibodies and the CD4+ T Cell MicroBead Cocktail. The magnetically labeled non-target T cells were depleted by retaining them on a MACS® Column in the magnetic field of a MACS Separator, while the unlabeled T helper cells passed through the column. The CD4+T were then lysed immediately in RLT lysis buffer (Qiagen Inc) with beta mercaptoethanol and stored at -80C. RNA & DNA were extracted simultaneously from the lysed CD4+T using the AllPrep protocol (Qiagen Inc). Library preparation was performed using TruSeq total RNA Ribo Zero kit (Illumina Inc) and paired end sequencing was performed at the University of Colorado genomics core on the HISEQ 4000, yielding 80 million paired end reads with read length of 151bp. BAGS CD4+T RNAseq data is available at [GEO accession].

*QC and Pre-Processing.* CoCo[10] was used to adjust the Ensembl annotation file for alignment and assembly. FastQC was used to detect technical issues, then adapter trimming was performed using BBDuk. FastQC was then run again to confirm trimming was successful. HISAT2 was used for alignment to GRCh38 using the default options. Sorting and flagging of reads was performed using SAMtools[11] prior to transcript assembly with StringTie[12] and applying the CoCo correct_count function using the corrected Ensembl annotation file to account for overlapping genes and multi-mapping reads. Raw counts were converted to transcripts per million (TPM) values for each gene and each sample.

Within-sample TPM distributions were used to detect and exclude samples where data generation was not successful. Additional sample-level metrics such as ribosomal contamination were calculated using Picard CollectRnaSeqMetrics. Transcripts with inadequate variability for detection of differential expression were filtered using the DESeq2[13] theta value. The DESeq2 varianceStabilizingTransformation function was used to transform the count data (normalized by division by the size factors or normalization factors), to be approximately homoscedastic and normalized with respect to library size.

Technical variability present in the cleaned and transformed TPM values due to sequencing batch was removed with the removeBatchEffect function in limma[14]. The transformed, batch-adjusted TPM values were used in prediction model building.

### Global Genetic Ancestry Deconvolution

Global ancestry proportions were estimated using ADMIXTURE 1.3[15]. K reference populations were selected separately by analysis set either by running cross validation on the founders in the population (CAAPA included in nasal epithelium model building, K=3)[1], or by prior knowledge of the population structure (BAGS included in CD4+T model building, K=2; BAGS-WGS included in CAAPA TWAS, K=2). The median global YRI genetic ancestry proportion was 0.84 for the nasal epithelium prediction population and 0.88 for the CD4+T prediction population (**Figure E1, Table E1**). For the TWAS populations, the mean proportion YRI ancestry was 0.88 for BAGS-WGS and 0.72 (range: 0.34-0.88) across all CAAPA studies, as reported by Daya et al[1] (**Table E2**).

### Prediction Model Building

We built separate transcriptome prediction databases for nasal epithelium and CD4+T following established methodology[16]. First, we quantified latent factors that capture technical variability and unmeasured confounding in each RNAseq dataset (e.g. tissue) using probabilistic estimation of expression residuals (PEER)[17]. We performed linear regression to adjust each gene's expression values by the recommended number of PEER factors based on sample size, sex, age, asthma status, and the first genetic principal component for CD4+T, and based on sample size, sex, age, asthma status, batch, study site, Agilent RNA integrity number equivalent, GC content, and the first two genetic principal components for nasal epithelium. The expression residuals were retained for prediction modeling using elastic net regression to select the set of genetic variants that best predicted gene expression. Eligible predictors for each gene included cis-SNPs within +/-500 Kb of the gene start/stop. Model prediction accuracy was determined using a nested, 10-fold cross-validation procedure[16]. Briefly, the model randomly split the data into five folds, then for each fold removed that fold from the data and used the remining data to train the elastic net model with 10-fold cross-validation for tuning of the lambda parameter. With the trained model, expression was predicted on the hold out fold to determine model performance by comparing the correlation between predicted and observed expression values. The procedure was repeated for each hold-out fold and the model performance metrics calculated as the average across each train-test set. Predictors (SNPs) and weights were selected from a final elastic net model trained using all subjects. Genes were considered significantly predicted and saved into the database if the average Pearson correlation between predicted and observed gene expression was greater than 0.1 (equivalent to $R^2>0.01$) with an estimated $P<0.05$ and at least 1 SNP was used for building the model[16].

Of the 16,728 genes expressed in CD4+T, significant prediction models were obtained for 8,351 (49.9%). Of the 14,766 genes expressed in nasal epithelium, 10,296 (69.7%) significant prediction models were obtained. There were 12,818 genes predicted in both databases, with 5,829 of those genes (45.4%) predicted well in both CD4+T and nasal epithelium. There were 4,467 (34.8%) genes only identified in the nasal epithelium database, and 2,522 (19.7%) only identified in the CD4+T database. Of the 172 unique asthma and allergy candidate genes (see "Candidate Genes and Enrichment Analyses"), 162 were found in at least one of the novel or existing prediction databases (see "Prediction of Gene Expression"). Of the 162, 52 (30.2%) were predictable in CD4+T and 91 (52.9%) in nasal epithelium (**Figure E2**).

### Prediction of Gene Expression

Gene expression was predicted from SNP genotypes in the 14 CAAPA studies used for TWAS in a total of 53 tissues. We predicted gene expression for individuals in all studies using the novel tissue databases (CD4+T, nasal epithelium), all existing GTEx version 8 prediction databases[18] (N=49) trained in a primarily European-ancestry population (85% European American, 12% African American), and prediction databases trained in African-ancestry populations: monocytes from the Multi-Ethnic Study of Atherosclerosis (MESA)[19], and whole blood from the Genes-environments and Admixture in Latino Americans and Study and the Study of African Americans, Asthma, Genes, and Environments (GALA-SAGE)[20].

### Prediction Accuracy Evaluation

To assess the generalizability of the novel CD4+T and nasal epithelium gene expression models across ancestral populations, we used the publicly available RNAseq and genetic data from the GEUVADIS consortium[21]. GEUVADIS completed RNA sequencing of cell lines generated from samples collected by the 1000 Genomes Project in five populations: West African ancestry Yoruba in Ibadan, Nigeria (YRI), Utah residents with Northern and Western European ancestry (CEU), Toscani in Italia (TSI), British in England and Scotland (GBR), and Finnish in Finland (FIN). We used the novel transcriptome prediction databases to estimate CD4+T and nasal epithelium gene expression for YRI (N=86) and the four European ancestry populations: CEU (N=89), TSI (N=93), GBR (N=92), and FIN (N=95). There were 7,272 genes with both observed expression from GEUVADIS and predicted CD4+T expression and 9,117 genes with observed expression from GEUVADIS and predicted nasal epithelium expression.

We examined prediction accuracy overall, across quintiles of model-based prediction accuracy, and across ancestral populations within each quintile. Prediction accuracy was calculated as the square of the Pearson correlation between predicted and observed gene expression, and compared across model performance thresholds and ancestry groups. We tested whether accuracy differed between any ancestral groups (K=5) using a Kruskal-Wallis test.

**Transcriptome-Wide Association Study and Meta-Analysis**

We performed TWAS using PrediXcan and MultiXcan[22] frameworks to identify candidate genes for asthma. For genes with tissue-specific causal effects, tissue-specific PrediXcan has the greatest power[22]. MultiXcan improves power to detect target genes by integrating shared eQTL architecture across tissues in a multivariate regression framework[22]. The study population included 9,284 non-overlapping participants of the 14 CAAPA asthma genetics studies for which GWAS or WGS data and asthma case-control status were available (**Table E2**). Tissue-specific gene expression was predicted as described above ("Prediction of Gene Expression"). Genetic principle components and kinship were estimated within each study using the GENetic EStimation and Inference in Structured samples (GENESIS) pipeline[23]. We tested for association between tissue-specific gene expression in the four prediction databases trained in African-ancestry populations (CD4+T, nasal epithelium, monocytes[19], and whole blood[20]) and asthma within each study (N=14) using a linear mixed model adjusted for sex, age (except for ProAR where age was unavailable), and genetic principal components, the number of which was determined by the scree plot elbow from GENESIS for each study.

$$Gene\ \widehat{Expression} \sim Asthma + Age + Sex + PCs + Kinship$$

Due to study design differences across CAAPA studies (described in detail previously[1]), TWAS of predicted gene expression were performed separately by study. Test statistics for each gene were then combined across the 14 CAAPA studies using inverse-variance weighted meta-analysis, representing an overall (cross-study) estimate of the effect of gene expression on asthma. For each set of tissue-specific TWAS results, we combined associations across studies using inverse-variance weighted meta-analyses in the R package Metagen. We tested for heterogeneity across populations using Cochran's Q and the $I^2$ statistic (**Figure E3**). We corrected for multiple comparisons using Benjamini Hochberg adjustment and considered genes with adjusted $P<0.1$ significant. The summary statistics are provided in **Table E3**.

MultiXcan takes advantage of shared eQTL architecture across tissues to improve TWAS power by considering gene expression-asthma effects jointly across multiple tissues. Given that LD structure may affect TWAS performance and varies across ancestry, we implemented MultiXcan in the 14 CAAPA studies using all available prediction databases (GTEx N=49, monocytes[19], whole blood[20], and the novel nasal epithelium and CD4+T databases). We then combined the gene-level model p-value across all 14 studies using a weighted Fisher's test, correcting for multiple comparisons using a Benjamini-Hochberg, considering genes with adjusted $P<0.1$ to be significant. The summary statistics are provided in **Table E4**.

**Candidate Genes and Enrichment Analyses**

Asthma and allergy candidate genes were identified from the GWAS catalog and through a literature search, as described in detail in Kanchan et al[24]. Gene symbols were mapped to Ensembl gene IDs using biomaRt, and all lookups and comparisons were completed using Ensembl gene ID. We used FUMA[25] (version 1.5.2) to perform all enrichment analyses.

GENE2FUNC from FUMA performs functional mapping, annotation, and enrichment analyses from input SNP or gene lists. To identify novel contributions to gene expression prediction from our CD4+T and nasal epithelium databases, we input the list of Ensembl gene IDs with prediction models uniquely identified in our novel tissues and not present in any existing external database. GENE2FUNC was run with a background set of all genes, Ensembl v110 and GTEx version 8 data sets, at least two or more minimum overlapping genes within gene sets, including the MHC region, and using Benjamini-Hochberg correction. Enriched Gene Ontology (GO) terms or pathways with false discovery rate (FDR) adjusted $P<0.05$ were considered significant.

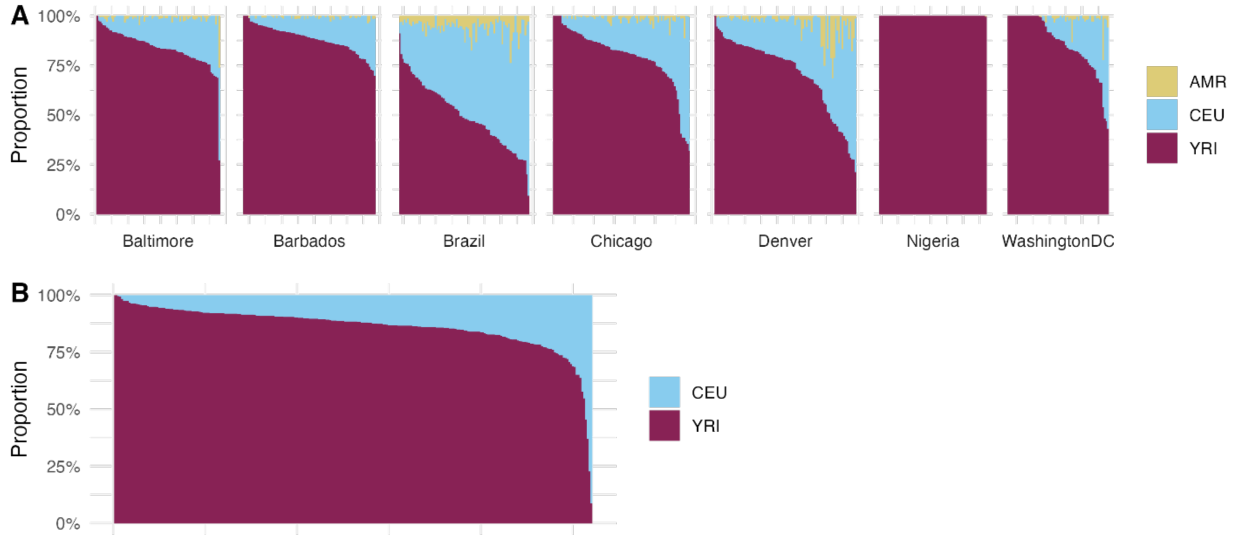


**Figure E1.** Global ancestry proportions for individuals included in training of new prediction databases: A) CAAPA individuals included in nasal epithelium model building, estimated using ADMIXTURE with K=3, and B) BAGS individuals included in CD4+T model building, estimated using ADMIXTURE with K=2. *AMR = Native American (selected from Mao et al*[26]*), CEU = Utah residents, YRI = Yoruba.*

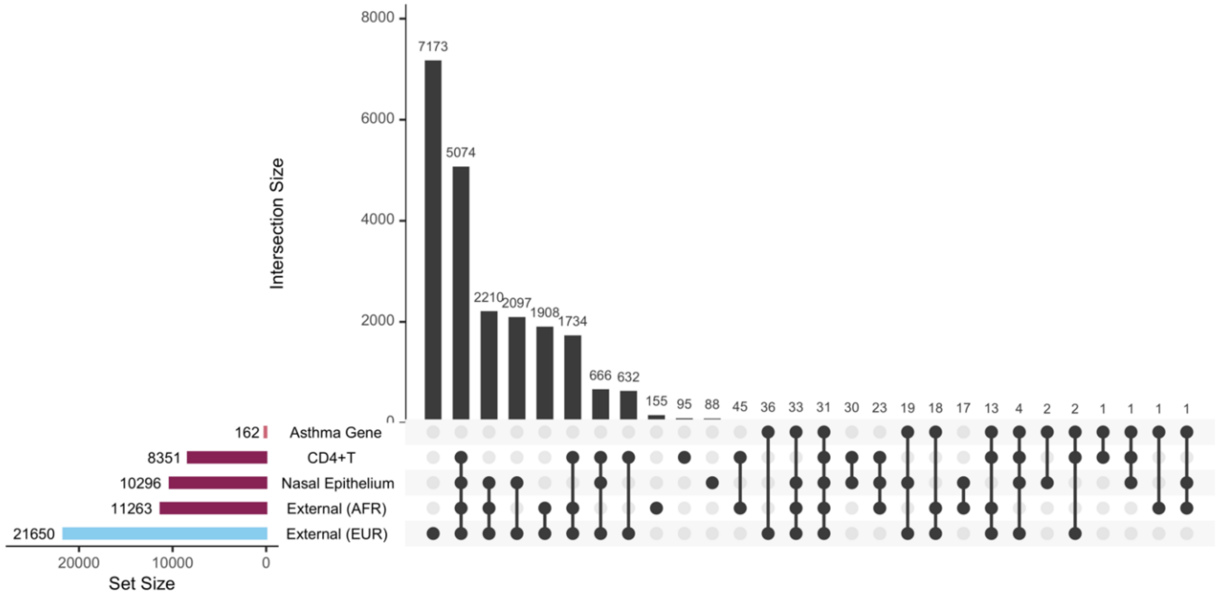


**Figure E2.** Upset plot of novel database gene overlap with external African- and European-ancestry trained tissue databases (total gene N=22,109) and well as candidate asthma genes.

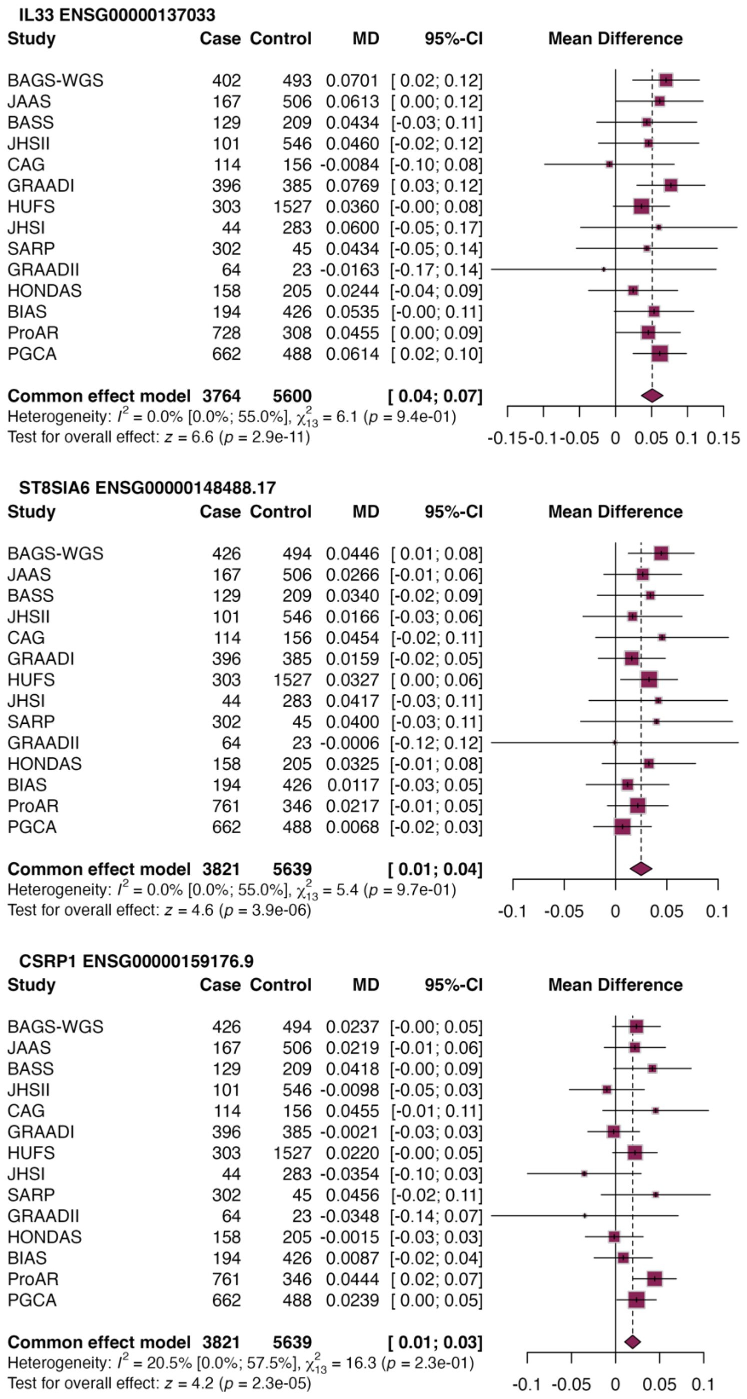


**Figure E3.** Forest plots of three genes identified as significant in meta-analysis from tissue-specific TWAS for asthma: *IL33* in nasal epithelium, *ST8SIA6* in whole blood, and *CSRP1* in monocytes. Studies are ordered by decreasing percent African ancestry (see Table E2), and the unadjusted p-value is shown.

### REFERENCES

1. Daya M, Rafaels N, Brunetti TM, et al. Association study in African-admixed populations across the Americas recapitulates asthma risk loci in non-African populations. *Nat Commun*. Feb 20 2019;10(1):880. doi:10.1038/s41467-019-08469-7

2. Szczesny B, Boorgula MP, Chavan S, et al. Multi-omics in nasal epithelium reveals three axes of dysregulation for asthma risk in the African Diaspora populations. *Nat Commun*. May 28 2024;15(1):4546. doi:10.1038/s41467-024-48507-7

3. Barnes KC, Freidhoff LR, Horowitz EM, et al. Physician-derived asthma diagnoses made on the basis of questionnaire data are in good agreement with interview-based diagnoses and are not affected by objective tests. *J Allergy Clin Immunol*. Oct 1999;104(4 Pt 1):791-6. doi:10.1016/s0091-6749(99)70289-7

4. Zambelli-Weiner A, Ehrlich E, Stockton ML, et al. Evaluation of the CD14/-260 polymorphism and house dust endotoxin exposure in the Barbados Asthma Genetics Study. *J Allergy Clin Immunol*. Jun 2005;115(6):1203-9. doi:10.1016/j.jaci.2005.03.001

5. Shaun Purcell CC. PLINK1.9. www.cog-genomics.org/plink/1.9/

6. Andrews S. FastQC: A Quality Control Tool for High Throughput Sequence Data. http://www.bioinformatics.babraham.ac.uk/projects/fastqc/

7. Picard Tools. Broad Institute. http://broadinstitute.github.io/picard

8. Bushnell B. BBMap. https://sourceforge.net/projects/bbmap/

9. Kim D, Paggi JM, Park C, Bennett C, Salzberg SL. Graph-based genome alignment and genotyping with HISAT2 and HISAT-genotype. *Nat Biotechnol*. Aug 2019;37(8):907-915. doi:10.1038/s41587-019-0201-4

10. Deschamps-Francoeur G, Boivin V, Abou Elela S, Scott MS. CoCo: RNA-seq read assignment correction for nested genes and multimapped reads. *Bioinformatics*. Dec 1 2019;35(23):5039-5047. doi:10.1093/bioinformatics/btz433

11. Danecek P, Bonfield JK, Liddle J, et al. Twelve years of SAMtools and BCFtools. *Gigascience*. Feb 16 2021;10(2)doi:10.1093/gigascience/giab008

12. StringTie. Johns Hopkins University Center for Computational Biology. https://ccb.jhu.edu/software/stringtie/

13. Love MI, Huber W, Anders S. Moderated estimation of fold change and dispersion for RNA-seq data with DESeq2. *Genome Biol*. 2014;15(12):550. doi:10.1186/s13059-014-0550-8

14. Ritchie ME, Phipson B, Wu D, et al. limma powers differential expression analyses for RNA-sequencing and microarray studies. *Nucleic Acids Res*. Apr 20 2015;43(7):e47. doi:10.1093/nar/gkv007

15. Alexander DH, Novembre J, Lange K. Fast model-based estimation of ancestry in unrelated individuals. *Genome Res*. Sep 2009;19(9):1655-64. doi:10.1101/gr.094052.109

16. Gamazon ER, Wheeler HE, Shah KP, et al. A gene-based association method for mapping traits using reference transcriptome data. *Nat Genet*. Sep 2015;47(9):1091-8. doi:10.1038/ng.3367

17. Stegle O, Parts L, Piipari M, Winn J, Durbin R. Using probabilistic estimation of expression residuals (PEER) to obtain increased power and interpretability of gene expression analyses. *Nat Protoc*. Feb 16 2012;7(3):500-7. doi:10.1038/nprot.2011.457

18. Barbeira AN, Bonazzola R, Gamazon ER, et al. Exploiting the GTEx resources to decipher the mechanisms at GWAS loci. *Genome Biol*. Jan 26 2021;22(1):49. doi:10.1186/s13059-020-02252-4

19. Mogil LS, Andaleon A, Badalamenti A, et al. Genetic architecture of gene expression traits across diverse populations. *PLoS Genet*. Aug 2018;14(8):e1007586. doi:10.1371/journal.pgen.1007586

20. Kachuri L, Mak ACY, Hu D, et al. Gene expression in African Americans, Puerto Ricans and Mexican Americans reveals ancestry-specific patterns of genetic architecture. *Nature Genetics*. 2023/05/25 2023;doi:10.1038/s41588-023-01377-z

21. Lappalainen T, Sammeth M, Friedländer MR, et al. Transcriptome and genome sequencing uncovers functional variation in humans. *Nature*. Sep 26 2013;501(7468):506-11. doi:10.1038/nature12531

22. Barbeira AN, Pividori M, Zheng J, Wheeler HE, Nicolae DL, Im HK. Integrating predicted transcriptome from multiple tissues improves association detection. *PLoS Genet*. Jan 2019;15(1):e1007889. doi:10.1371/journal.pgen.1007889

23. Gogarten SM, Sofer T, Chen H, et al. Genetic association testing using the GENESIS R/Bioconductor package. *Bioinformatics*. 2019;35(24):5346-5348. doi:10.1093/bioinformatics/btz567

24. Kanchan K, Clay S, Irizar H, Bunyavanich S, Mathias RA. Current insights into the genetics of food allergy. *J Allergy Clin Immunol*. Jan 2021;147(1):15-28. doi:10.1016/j.jaci.2020.10.039

25. Watanabe K, Taskesen E, van Bochoven A, Posthuma D. Functional mapping and annotation of genetic associations with FUMA. *Nat Commun*. Nov 28 2017;8(1):1826. doi:10.1038/s41467-017-01261-5

26. Mao X, Bigham AW, Mei R, et al. A genomewide admixture mapping panel for Hispanic/Latino populations. *Am J Hum Genet*. Jun 2007;80(6):1171-8. doi:10.1086/518564
